# Spatial and temporal trends in social vulnerability and COVID-19 incidence and death rates in the United States

**DOI:** 10.1101/2020.09.09.20191643

**Authors:** Brian Neelon, Fedelis Mutiso, Noel T Mueller, John L Pearce, Sara E Benjamin-Neelon

## Abstract

**Background:** Emerging evidence suggests that socially vulnerable communities are at higher risk for coronavirus disease 2019 (COVID-19) outbreaks in the United States. However, no prior studies have examined temporal trends and differential effects of social vulnerability on COVID-19 incidence and death rates. The purpose of this study was to examine temporal trends among counties with high and low social vulnerability and to quantify disparities in these trends over time. We hypothesized that highly vulnerable counties would have higher incidence and death rates compared to less vulnerable counties and that this disparity would widen as the pandemic progressed.

**Methods:** We conducted a retrospective longitudinal analysis examining COVID-19 incidence and death rates from March 1 to August 31, 2020 for each county in the US. We obtained daily COVID-19 incident case and death data from USAFacts and the Johns Hopkins Center for Systems Science and Engineering. We classified counties using the Social Vulnerability Index (SVI), a percentile-based measure from the Centers for Disease Control and Prevention in which higher scores represent more vulnerability. Using a Bayesian hierarchical negative binomial model, we estimated daily risk ratios (RRs) comparing counties in the first (lower) and fourth (upper) SVI quartiles. We adjusted for percentage of the county designated as rural, percentage in poor or fair health, percentage of adult smokers, county average daily fine particulate matter (PM2.5), percentage of primary care physicians per 100,000 residents, and the proportion tested for COVID-19 in the state.

**Results:** In unadjusted analyses, we found that for most of March 2020, counties in the upper SVI quartile had significantly fewer cases per 100,000 than lower SVI quartile counties. However, on March 30, we observed a “crossover effect” in which the RR became significantly greater than 1.00 (RR = 1.10, 95% PI: 1.03, 1.18), indicating that the most vulnerable counties had, on average, higher COVID-19 incidence rates compared to least vulnerable counties. Upper SVI quartile counties had higher death rates on average starting on March 30 (RR = 1.17, 95% PI: 1.01,1.36). The death rate RR achieved a maximum value on July 29 (RR = 3.22, 95% PI: 2.91, 3.58), indicating that most vulnerable counties had, on average, 3.22 times more deaths per million than the least vulnerable counties. However, by late August, the lower quartile started to catch up to the upper quartile. In adjusted models, the RRs were attenuated for both incidence cases and deaths, indicating that the adjustment variables partially explained the associations. We also found positive associations between COVID-19 cases and deaths and percentage of the county designated as rural, percentage of resident in fair or poor health, and average daily PM_2.5_.

**Conclusions:** Results indicate that the impact of COVID-19 is not static but can migrate from less vulnerable counties to more vulnerable counties over time. This highlights the importance of protecting vulnerable populations as the pandemic unfolds.

## Introduction

Severe acute respiratory syndrome coronavirus 2 (SARS-CoV-2), the cause of coronavirus disease 2019 (COVID-19), has created a global public health crisis since its onset in late 2019. As of September 1, 2020, there have been over 6 million confirmed COVID-19 cases and over 183,000 related deaths in the United States (US) alone [1]. Emerging evidence indicates that the pandemic disproportionately affects people of color, older individuals, and those of lower socioeconomic status [2–7]. Recent data suggest that African Americans are contracting COVID-19 at higher rates and are more likely to die from the virus [6, 8]. Two studies also reported that COVID-19 infection rates are greater in US counties and in states with high Latinx populations and monolingual Spanish speakers [4, 7]. Further, earlier studies from China found that older age was associated with an increased risk of death among those infected with COVID-19 [5, 9]. Older age was also associated with COVID-related hospitalizations in New York City [10]. Underlying health conditions and comorbidities may partially explain these associations [5], but do not fully account for the disproportionate burden. Recent studies suggest that social determinants of health and community contextual factors contribute to these disparities, and that socially vulnerable communities are at highest risk for COVID-19 outbreaks [6, 11–13].

Protecting vulnerable populations is critically important during the COVID-19 pandemic, as these groups are generally at higher risk for adverse health outcomes [14, 15]. Hurst et al. define vulnerability as an identifiably elevated risk of incurring greater wrong or harm [16]. One type of vulnerability – social vulnerability – has been used by the Centers for Disease Control and Prevention (CDC) to identify communities most at risk when faced with adverse events that may impact health, such as natural disasters or disease outbreaks. The CDC developed the social vulnerability index (SVI) to assist federal, state, and local governments in targeting and mobilizing resources for at-risk counties in response to adverse events.

Recent studies have demonstrated the importance of considering social vulnerability in both COVID-19 cases and deaths, although the findings have been somewhat inconsistent [17–19]. Karaye et al. examined associations between the SVI and cumulative COVID-19 cases on May 12, 2020 [17]. They found that SVI total score was associated with increased rates of COVID-19. However, the authors found no association when they examined six states with high testing rates. Khazanchi and colleagues conducted an analysis of COVID-19 cases and deaths through April 19, 2020, and found that those living in the most vulnerable counties (highest SVI) had greater risk of infection and death [19]. Nayak et al. examined associations between the SVI and COVID-19 incidence and case fatalities through April 4, 2020, and found a significant association between social vulnerability and case fatality but not incident cases [18]. Notably, all three studies were cross-sectional and conducted at different time points early in the pandemic, which might contribute to the inconsistent findings. In fact, to date, no prior studies have examined longitudinal trends in social vulnerability and COVID-19 incidence and death rates in an effort to determine how these relationships change over time. Therefore, the purpose of this study was to examine temporal trends among counties with high and low social vulnerability and to quantify disparities in these trends over time.

## Methods

### Overview

We conducted a retrospective longitudinal analysis examining COVID-19 incidence and death rates from March 1, 2020 to August 31, 2020 for each of the 3,142 US county and county equivalents based on their unique Federal Information Processing Series (FIPS) codes [20, 21]. Specifically, we modeled the temporal trend in daily incidence and death rates for each county and assessed differential risks by county-level social vulnerability. We hypothesized that highly vulnerable counties would have higher incidence and death rates compared to less vulnerable counties and that this disparity would widen over time. The Institutional Review Boards at the Medical University of South Carolina and Johns Hopkins Bloomberg School of Public Health deemed this research exempt from review.

### COVID-19 Incident Cases and Deaths

We obtained daily COVID-19 incident case and death data from USAFacts [22] and the Johns Hopkins Center for Systems Science and Engineering [23]. Because Johns Hopkins aggregates data for some counties (e.g., the five boroughs of New York) [24], we opted to use the USAFacts data in our primary analysis, and conducted a sensitivity analysis using Johns Hopkins data. For both data sources, we downloaded daily incident case and death counts from March 1 to August 31, 2020. We obtained county population data from the 2019 population datafile compiled by the US Census Bureau [25].

### Social Vulnerability Index

We used publicly available data from the CDC’s Agency for Toxic Substances and Disease Registry to classify counties using SVI [26]. The SVI is a percentile-based measure of social vulnerability, or the resilience of communities to address stressors to health related to external hazards (e.g., natural disasters or disease outbreaks) [27]. The Geospatial Research, Analysis & Services Program within the Agency for Toxic Substances and Disease Registry created the SVI database to help public health officials identify communities that will most likely need support and resources during and after a hazardous event like a pandemic [26]. The overall index and each theme is scored from 0 to 1, with higher scores indicating greater vulnerability [26, 27]. The index was constructed using data from 15 variables from the US Census Bureau. A percentile rank was calculated for each of these variables and grouped among four themes of SVI that measure various aspects of vulnerability – these include Socioeconomic Status, Household Composition, Race/Ethnicity/Language, and Housing/Transportation [26, 27].

The Socioeconomic Status theme is composed of percentile rank data for the following variables: percentage below poverty, percentage unemployed, per capita income, and percentage with no high school diploma. For Household Composition, the variables include percentage age 65 years and older, percentage age 17 years or younger, percentage age 5 years or older with a disability, and percentage of single-parent households. The Race/Ethnicity/Language theme encompasses percentage minority and percentage who speaks English “less than well”. Finally, the Housing/Transportation theme includes data for the percentage of multiunit structures, percentage of mobile homes, percentage crowding, percentage having no vehicle, and percentage of group quarters.

For our analyses, we downloaded the 2018 county-level SVI data (the most recent available) for all 3,142 counties. One county was missing SVI data; for this county, we imputed SVI data using the national average.

### Adjustment Variables

We conducted both unadjusted and adjusted analyses for this study. For the adjusted analyses, we selected variables unrelated to the components of SVI that could explain the differential impact of COVID-19 on upper and lower SVI counties. These variables were chosen *a priori* based on previously reported associations with COVID-19 incidence and deaths [17–19, 28–31]. We obtained several health and environmental factors from the Robert Wood Johnson Foundation’s 2019 County Health Rankings & Roadmaps: Rankings Data & Documentation [32]. These included the percentage of each county designated as rural, the percentage of residents in poor or fair health, the percentage of adult smokers in the county, the average daily PM2.5 for each county, and the number of primary care physicians per 100,000 in each county. We also controlled for the cumulative proportion of COVID-19 Viral (RT-PCR) tests performed in each state through August 31, 2020, which we obtained from the CDC Covid Data Tracker [1] (county-level data are not currently available). We converted the number tested to a proportion by dividing the number of tests by the state population sizes, which we obtained from the US Census Bureau’s population estimate dataset [33].

### Statistical Analysis

We first conducted an unadjusted analysis to compare trends across high- and low-SVI counties; we then performed an adjusted analysis to determine whether the results changed substantially after controlling for potential confounders. For both analyses, we fit Bayesian hierarchical negative binomial models with daily incident cases and daily deaths for each county as the outcomes. The models included penalized cubic Bsplines for both the fixed and random (i.e., county-specific) temporal effects, with knots placed every two weeks over the study period (15 total). The models also included county population as an offset on the log scale to convert the case and death counts to population-adjusted rates.

To avoid overfitting the temporal splines, we assigned ridging priors to the fixed and county-specific spline coefficients – i.e., independent, mean-zero normal distributions with shared inverse gamma variances [34]. We assigned a gamma prior to the negative binomial dispersion parameter. We developed an efficient data-augmented Gibbs sampler to aid posterior computation [35, 36]. For both the incidence case and death rate models, we ran the Gibbs sampler for 2,500 iterations with a burn-in 500 to ensure convergence. In sensitivity analyses, we increased the number knots to 30 and found no appreciable difference in the results.

To report results, we compared counties in the top or upper SVI quartile (most vulnerable) to those in bottom or lower SVI quartile (least vulnerable). For both quartiles, we graphed the posterior mean incidence and death rate trends along with their 95% posterior intervals (PIs). We also reported risk ratios (RRs) and 95% PIs comparing the upper and lower quartiles on each day for the overall SVI and its themes. Additionally, we reported posterior mean trends for select counties with differing SVI profiles.

For comparison, we refit the models controlling for potential confounders listed above. We assigned weakly informative normal priors to the corresponding regression parameters. We graphed the incidence and death rate trends, as well RRs, for the reference covariate group in the adjusted analyses. We also reported posterior RRs and 95% PIs for the adjustment variables. We conducted all analyses using R software version 3.6 (R Core Team 2019, R: A language and environment for statistical computing, R Foundation for Statistical Computing, Vienna, Austria).

## Results

### Unadjusted Analysis

#### Overall SVI

The final analytic sample comprised 578,128 observations (3,142 counties × 184 study days). There were 786 counties in each of the upper and lower SVI quartiles. Figure 1A presents the per capita incidence trends (expressed as cases per 100,000) for the upper and lower quartiles of SVI from the unadjusted analysis. For counties in the upper quartile, the average incidence increased steadily from March 1 (estimated 0.002 cases per 100,000; 95% PI: 0.001, 0.004) to April 25 (8.04 cases per 100,000; 95% PI: 7.67, 8.47). The incidence leveled off from April 26 to June 4 (8.97 cases per 100,000; 95% PI: 8.70, 9.25) before a precipitous increase through July 23 (31.26 cases per 100,000; 95% PI: 30.62, 31.83). The incident cases declined thereafter, before a final uptick in late August (23.10 cases per 100,000 on August 31; 95% PI: 21.47, 25.12). The lower quartile exhibited a similar but less pronounced trend: there was a modest increase from March 1 (0.002 cases per 100,000; 95% PI: 0.002, 0.004) to April 1 (3.04 cases per 100,000; 95% PI: 2.91, 3.19) and a longer plateau lasting until June 16 (3.08 cases per 100,000; 95% PI: 2.97, 3.19). There was a modest increase from June 16 to July 20 (9.05 cases per 100,000; 95% PI: 8.78, 9.31) followed by a sharp increase in late August (17.37 cases per 100,000 on August 31; 95% PI: 16.00, 19.00).

**Figure 1.**
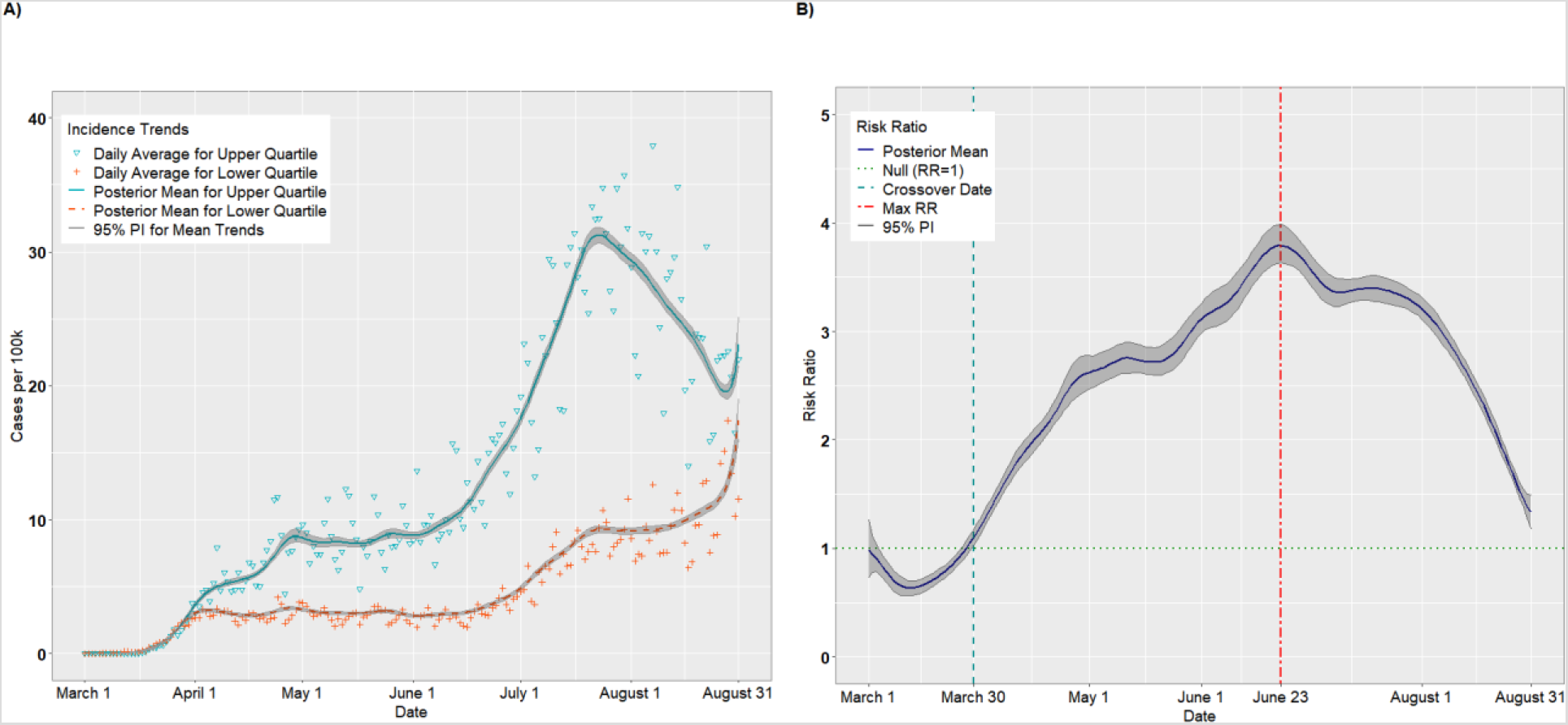
Per capita incidence (A) and risk ratios (B) comparing upper and lower quartiles of overall SVI

Figure 1B presents the posterior mean RRs comparing the upper and lower quartiles on each day. On March 1, the RR for incident cases was 0.99 (95% PI: 0.73, 1.26), suggesting that upper SVI quartile counties had, on average, fewer cases per 100,000 than lower SVI quartile counties, although this result did not statistically differ from 1.00. In fact, through March 27, the RRs were < 1.00. On March 12, for example, the RR comparing the upper to the lower quartile achieved its nadir at 0.63 (95% PI: 0.56, 0.71). However, on March 30, we observed a “crossover effect” in which the RR became significantly greater than 1.00, indicating that the more vulnerable counties had higher COVID-19 incidence on average compared to less vulnerable counties (March 30 RR = 1.10, 95% PI: 1.03, 1.18). The RRs increased steadily thereafter and achieved a maximum RR of 3.80 (95% PI: 3.63, 3.99) on June 23, then decreased steadily until August 31 (RR = 1.33, 95% PI: 1.18, 1.49). This suggests that the disparity in per capita cases between the upper and lower quartiles widened until late June, after which the lower quartile began to keep pace with the upper quartile.

Figure 2A presents per capita death trends (expressed as deaths per million) for the upper and lower quartiles of overall SVI. The death rates for both quartiles increased until April 26 before receding slightly in May and June. Beginning in early July, however, the mean death rate for the upper quartile increased steadily, achieving a maximum value on August 15 of 6.52 deaths per million (95% PI: 6.20, 6.88). Figure 2B presents the daily RRs comparing the upper and lower quartiles. Starting on March 30, the upper quartile had consistently higher death rates compared to the lower quartile (RR = 1.17, 95% PI: 1.01,1.36). The RRs increased until achieving a maximum value on July 29 (RR = 3.22, 95% PI: 2.91, 3.58) before tapering off in August (August 31 RR = 2.13, 95% PI: 1.72, 2.65).

**Figure 2.**
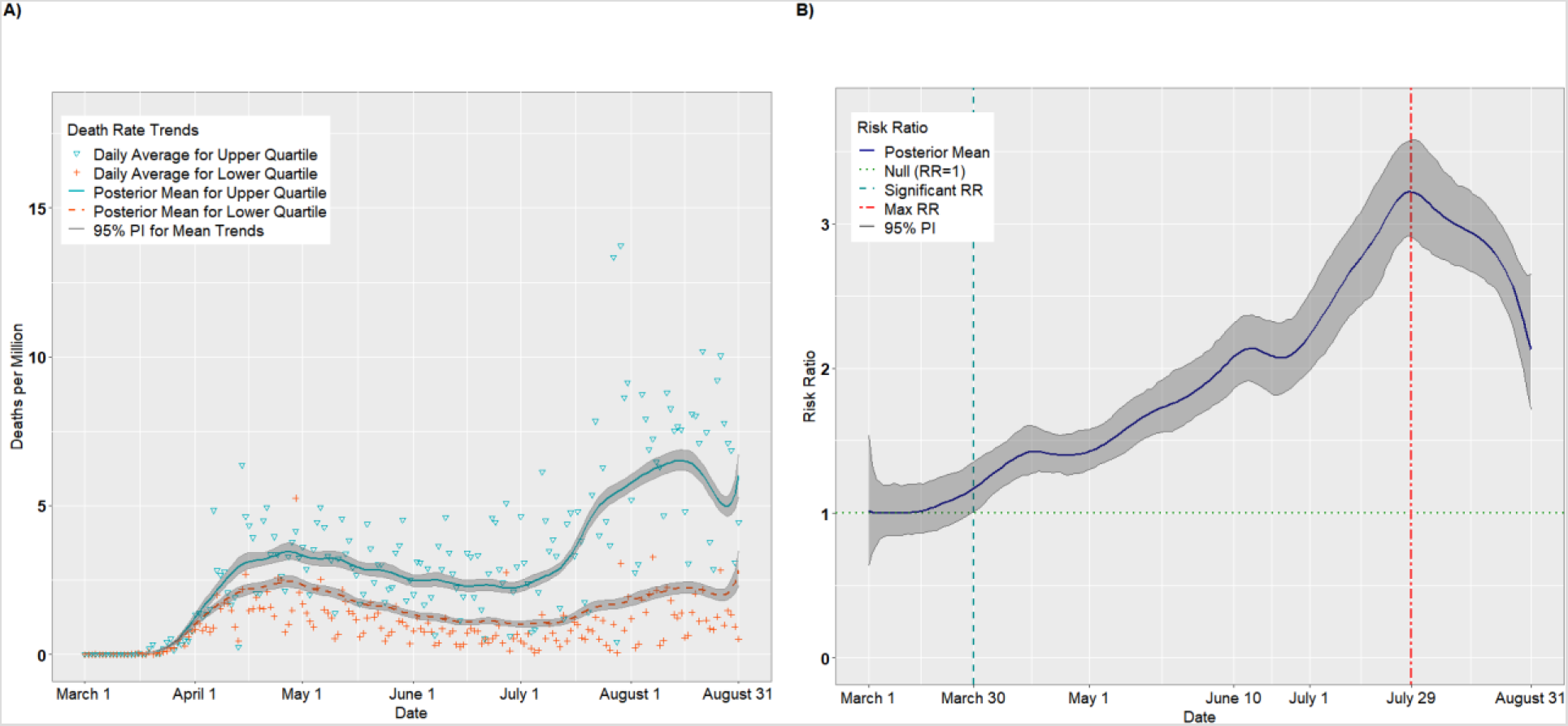
Per capita death rates (A) and risk ratios (B) comparing upper and lower quartiles of overall SVI

#### SVI Theme: Socioeconomic Status

Figures 3A-B and 4A-B present the temporal trends and RRs for incident cases and deaths, respectively, for the Socioeconomic Status theme. The trends were similar to those for overall SVI. According to Figure 3B, incident cases were higher for the lower Socioeconomic Status quartile from March 1 through April 3, with the lowest RR occurring on March 11 (RR = 0.52, 95% PI: 0.46, 0.58). Thus, on March 12, the most vulnerable counties had approximately half the incidence as the least vulnerable counties. As with overall SVI, there was a crossover effect on April 3 in which the RRs became significantly > 1.00. The RRs achieved a maximum of 2.94 (95% PI: 2.82, 3.06) on June 20 before a plateau in July. Starting in August, the RRs declined steadily as the per capita cases for the lower quartile began to catch up to the upper quartile (August 31 RR = 1.45, 95% PI: 1.32, 1.62). Likewise, as indicated in Figure 4B, the death rate was higher for the lower quartile than the upper quartile from March 1 through March 22, with the lowest RR occurring on March 6 (RR = 0.73, 95% PI: 0.55, 0.93). As with incident cases, The RRs became significantly positive on April 3 (RR = 1.15, 95% PI: 1.01, 1.31), and attained a maximum value of 2.97 (95% PI: 2.70 3.29) on July 30. Unlike with incident cases, however, the death rate disparity between upper and lower SES quartiles remained elevated through August 31 (RR = 2.35, 95% PI: 1.89, 2.88).

**Figure 3.**
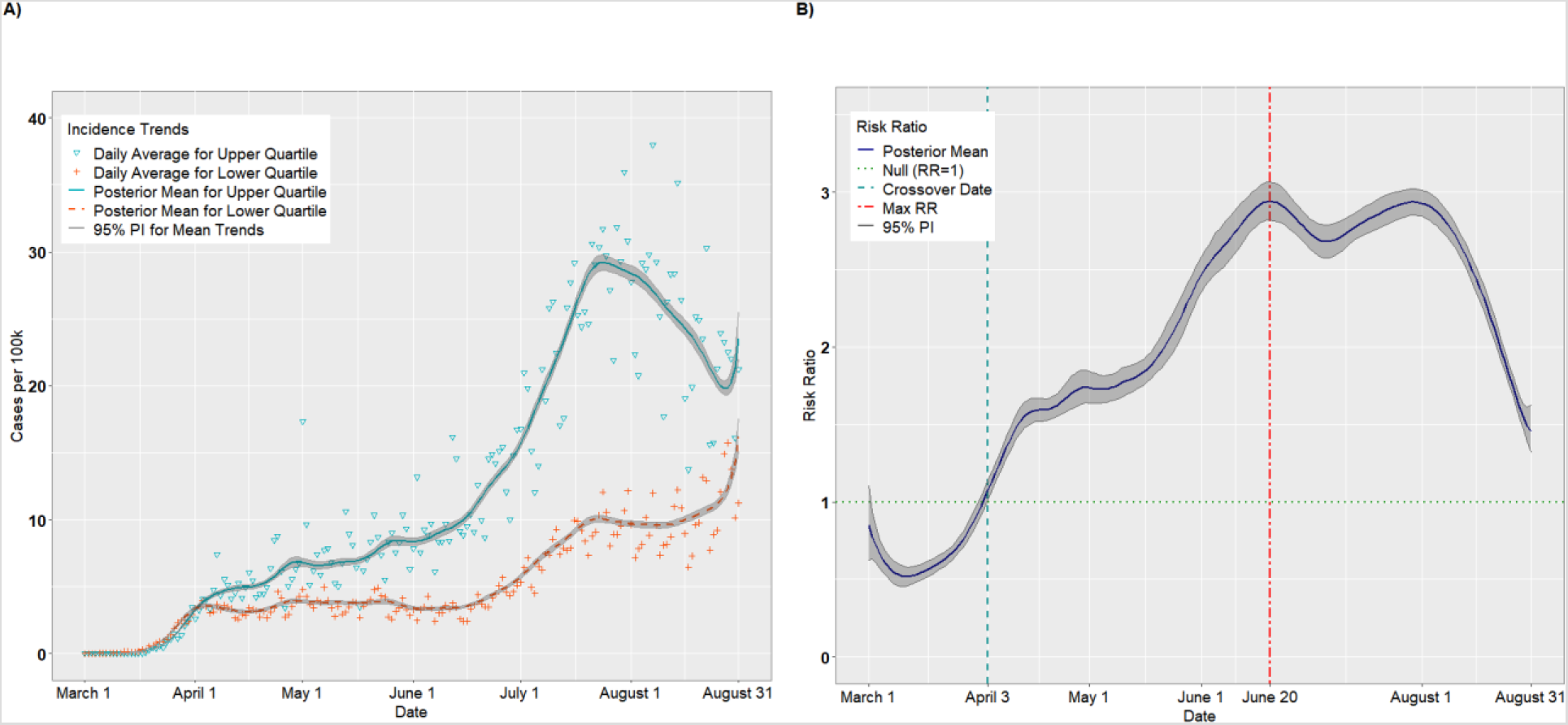
Per capita incidence (A) and risk ratios (B) comparing the upper to lower quartiles of the SVI Socioeconomic Status theme

**Figure 4.**
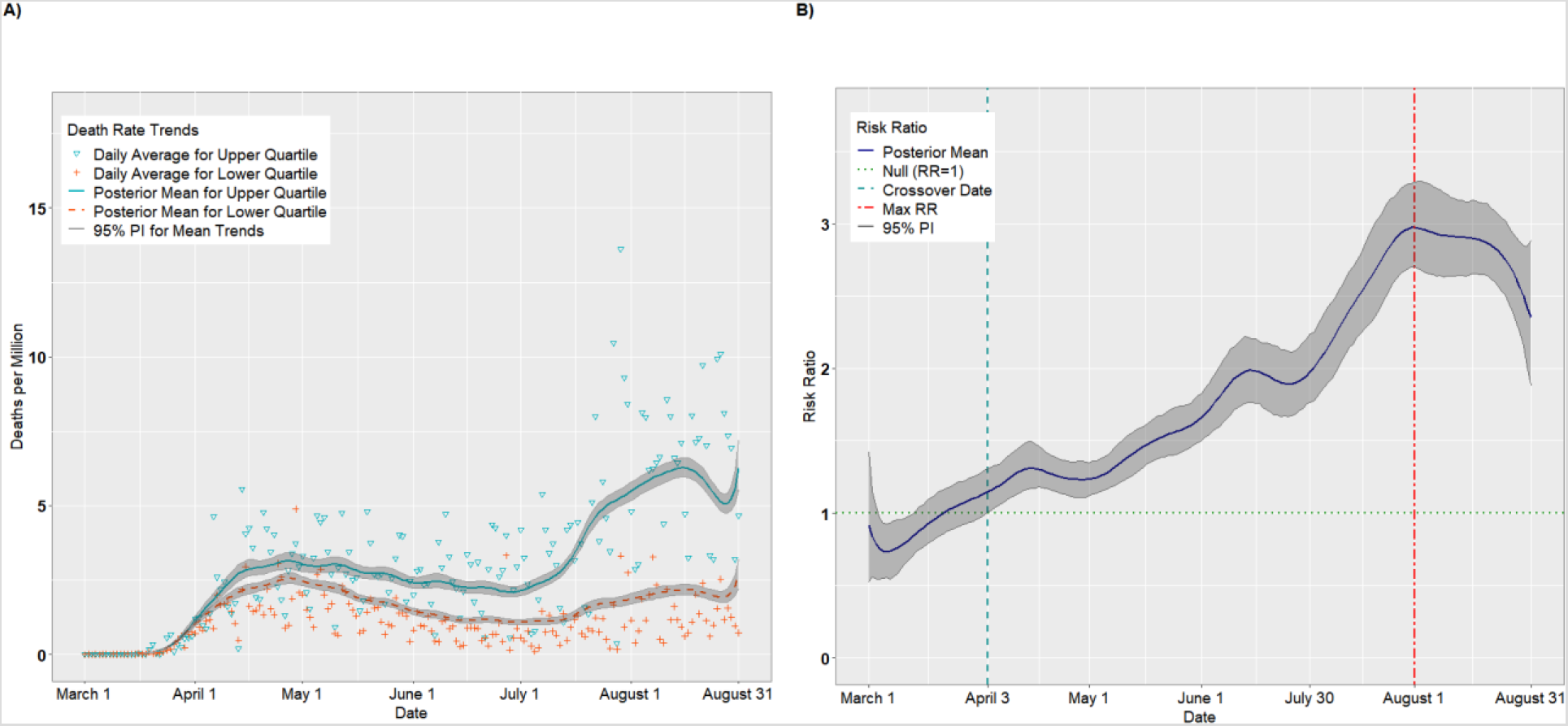
Per capita death rates (A) and risk ratios (B) comparing the upper to lower quartiles of the SVI Socioeconomic Status theme

#### SVI Theme: Household Composition

Figures 5A-B and 6A-B present the results for the Housing Composition theme. The crossover effect was significantly delayed for this theme, with the crossover dates occurring on May 16 for incident cases (Figure 5B) and on May 31 for deaths (Figure 6B). Thus, the pandemic appears to have disproportionately impacted the least vulnerable counties with respect to household composition for much of the early pandemic. However, these trends reversed by June. For incident cases, the daily RRs achieved a maximum of 2.05 (95% PI: 1.99, 2.10) on July 26 (Figure 5B) and then declined steadily. By August 31, there was a null association between upper and lower quartiles (RR = 1.00, 95% PI: 0.90, 1.10). For deaths (Figure 6B), the maximum RR of 2.34 (95% PI: 1.96, 2.33) was achieved August 16 and, unlike incident cases, remained above 2.0 through August 31 (RR = 2.10, 95% PI: 1.67, 2.57).

**Figure 5.**
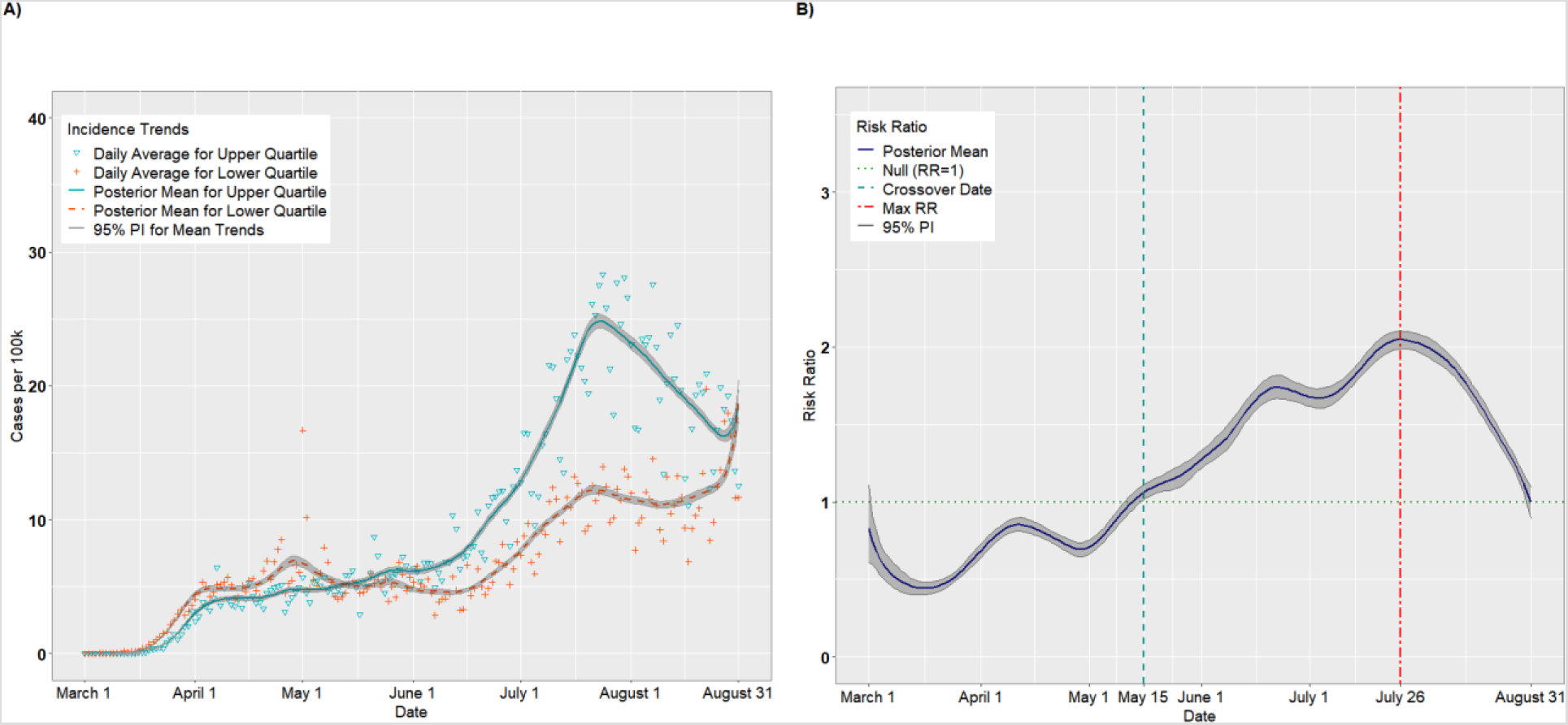
Per capita incidence (A) and risk ratios (B) comparing the upper to lower quartiles of the SVI Household Composition theme

**Figure 6.**
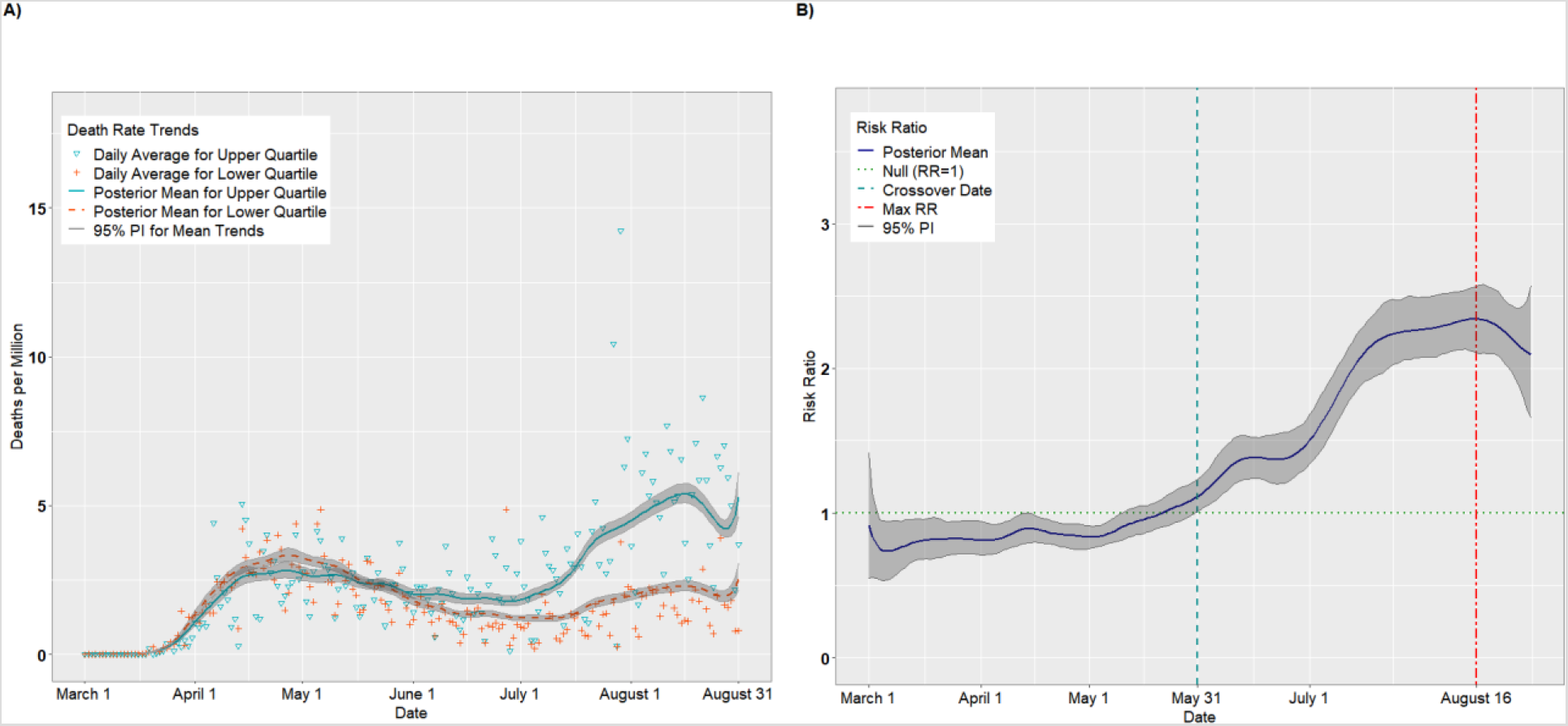
Per capita death rates (A) and risk ratios (B) comparing the upper to lower quartiles of the SVI Household Composition theme

#### SVI Theme: Race/Ethnicity/Language

Figures 7A-B and 8A-B present the results for the Race/Ethnicity/Language theme. Unlike the previous themes, vulnerable counties experienced higher incidence and death rates from the outset of the pandemic. In fact, the disparity between the upper and lower quartile was greatest for this theme, with a maximum incidence RR of 5.13 (95% PI: 4.84, 5.46) on May 2 and another local peak on June 24 (Figure 7B). For cases, the RRs declined steadily from late June into August, as the incidence for the lower quartile outpaced the upper quartile. By the end of August, there was no significant association between upper and lower quartiles with respect to incidence (RR = 0.92; 95% PI: 0.92, 1.02). In contrast, the death rate RRs (Figure 8B) hovered between 2 and 3 for most of the late spring and summer, before a decline in August.

**Figure 7.**
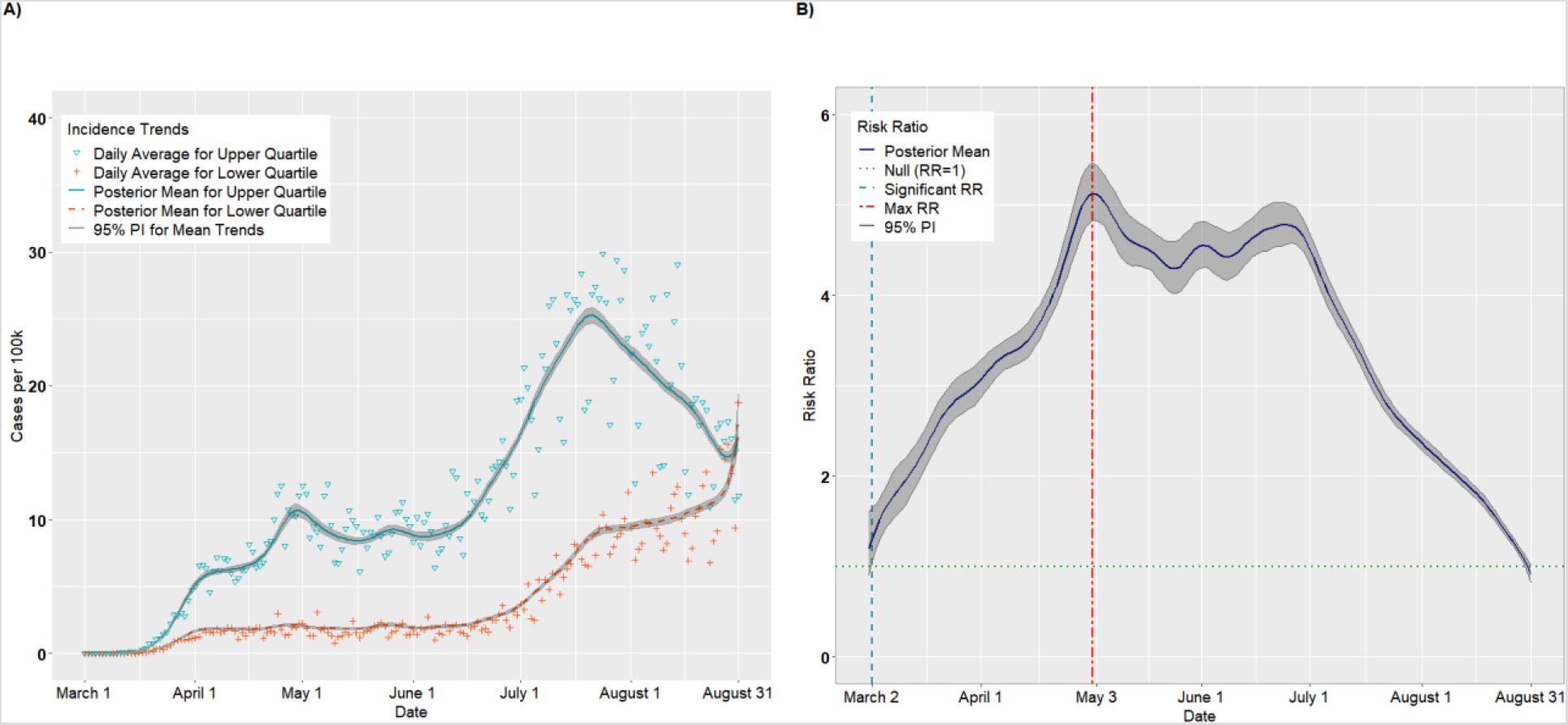
Per capita incidence (A) and risk ratios (B) comparing the upper to lower quartiles of the SVI Race/Ethnicity/Language theme

**Figure 8.**
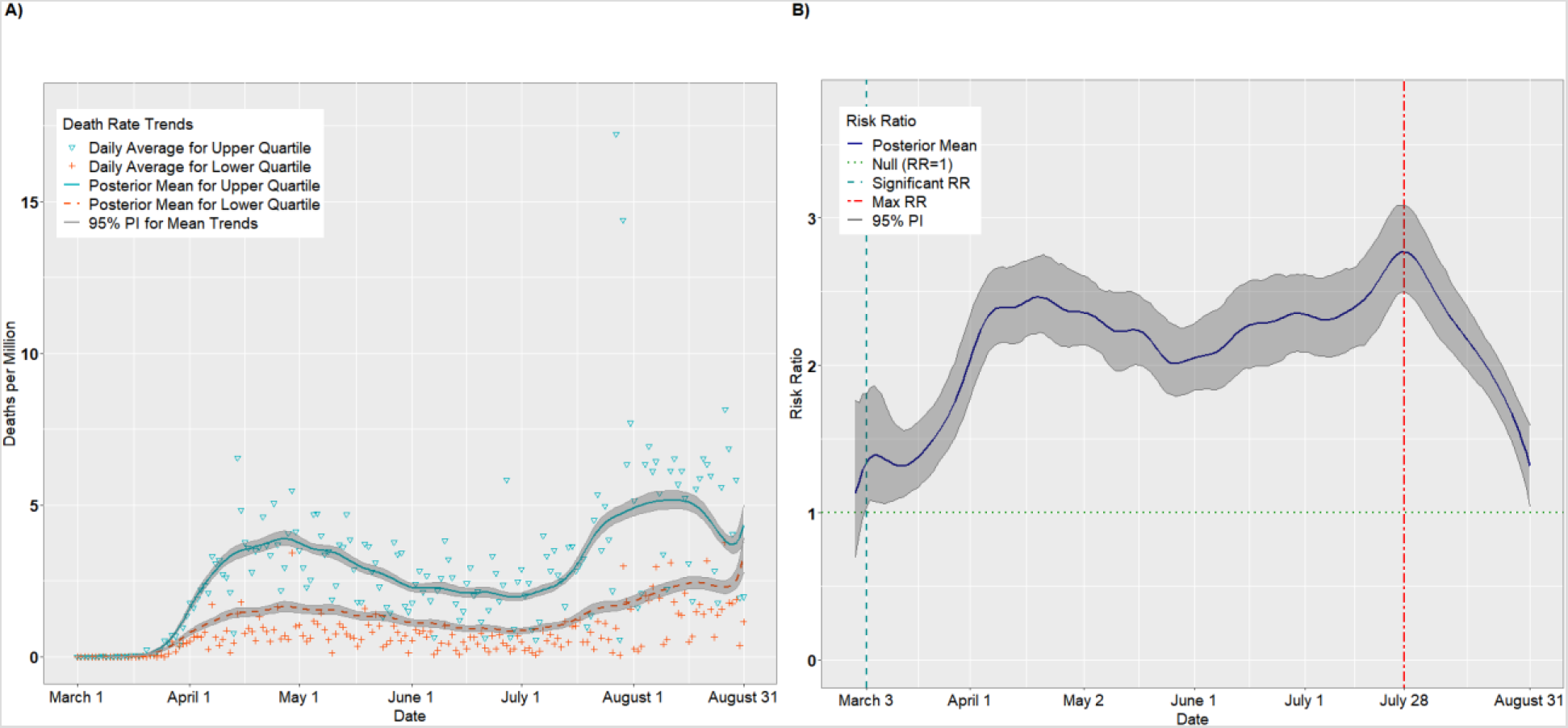
Per capita death rates (A) and risk ratios (B) comparing the upper to lower quartiles of the SVI Race/Ethnicity/Language theme

#### SVI Theme: Housing/Transportation

Figures 9A-B and 10A-B present the results for the Housing/Transportation theme. The incident case RRs (Figure 9B) remained significantly positive from March 4 (RR = 1.18, 95% PI: 1.01, 1.35) through August 31, achieving a maximum of 2.78 (95% PI: 2.66, 2.92) on April 28. The death rate RRs hovered around 2.00 for most of the study period, implying a uniform disparity between upper and lower quartile counties

**Figure 9.**
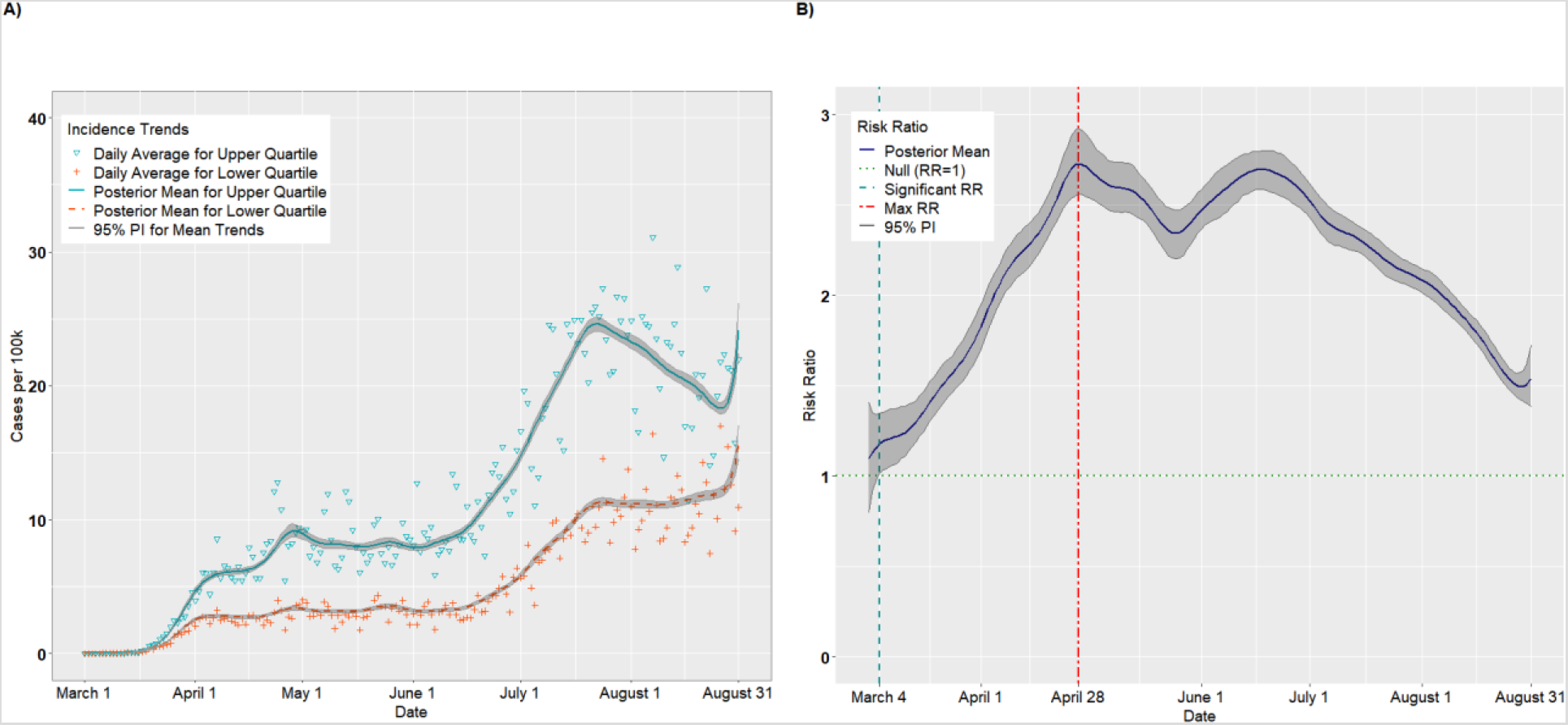
Per capita incidence (A) and risk ratios (B) comparing the upper to lower quartiles of the SVI Housing/Transportation theme

**Figure 10.**
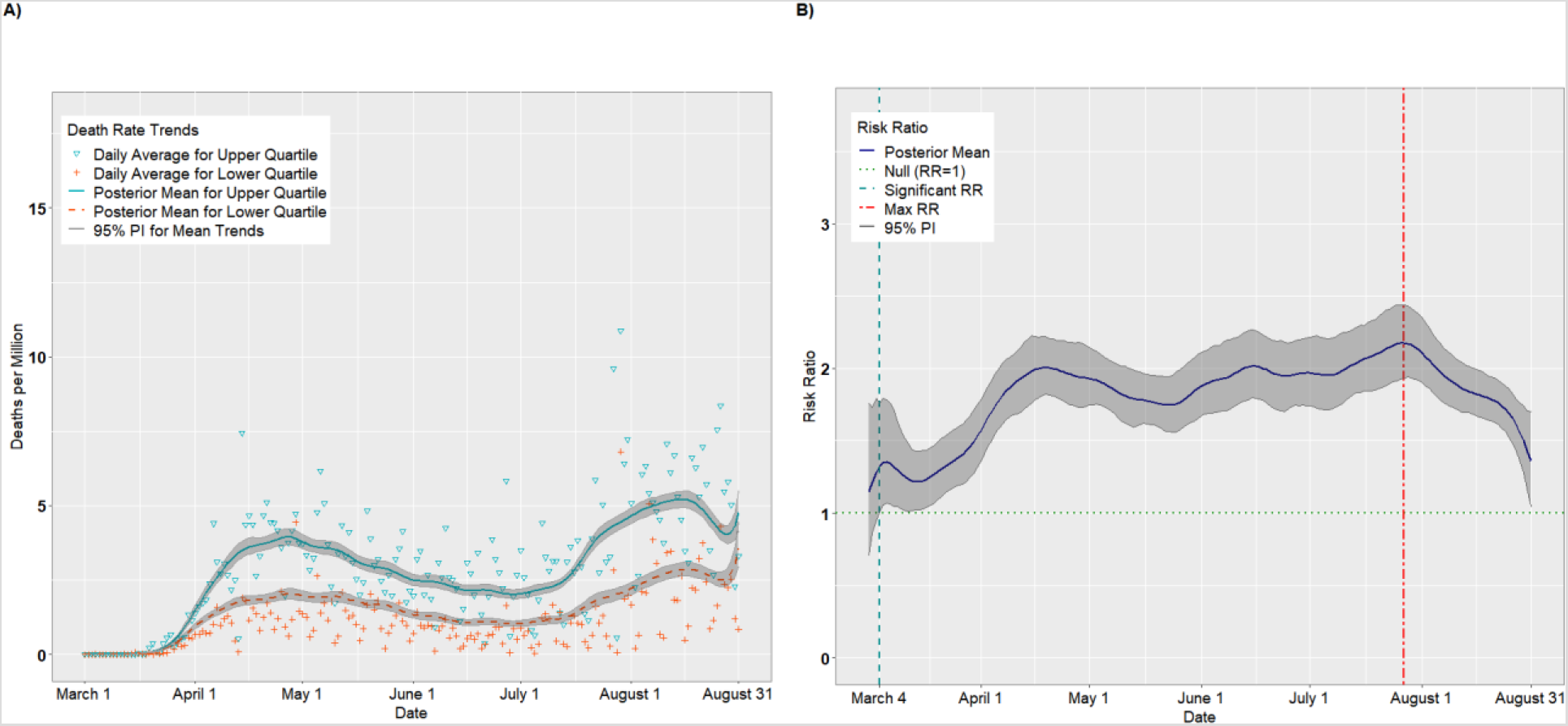
Per capita death rates (A) and risk ratios (B) comparing the upper to lower quartiles of the SVI Housing/Transportation theme

#### Illustrative Counties

Figures S1A-B in the Supporting Information present the incidence and death rate trends for Brooks County, Texas, the county with the highest overall SVI score of 1.00. As expected of high-SVI counties, the incidence and death rates remained low early in the pandemic, but began to escalate in July and early August. Figures S2A-B present analogous trends for Elbert County, Colorado, the county with the lowest overall SVI score of 0.00. The incidence and death rates remained relatively low throughout the pandemic, with a slight uptick in early August. This reflects the recent upward trend we observed in Figure S1A for counties in the lowest quartile.

Figures S3A-B and S4A-B present trends for two counties that illustrate the crossover effect we observed in Figure 1A, whereby lower quartile counties had higher average incidence than the upper quartile early in the pandemic. Figures S3A-B present results for Nassau County, New York, which has an overall SVI score of 0.24, placing it in the lower quartile. Here, both incidence cases and deaths spiked in early April before dissipating in May. In contrast, Figures S4A-B show the trends for Taylor County, Florida, an upper-quartile county with an overall SVI score of 0.90. As with Brooks County, the incidence and death rates were near zero until early August, when the rates increased substantially due to an outbreak at a local correctional facility [37].

#### Sensitivity Analysis Using Johns Hopkins Data

Sensitivity analysis using the Johns Hopkins data produced similar results to those we observed using USAFacts data. Figures S5A-B and S6A-B present the incidence and death rate trends for overall SVI. In all cases, the results were almost identical across the two data sources.

### Adjusted Analysis

Figures S7A-B and S8A-B present the overall-SVI incidence and death rate trends for the reference covariate group from the adjusted analyses. The incident case trends for overall SVI (Figures S7A-B) were similar to the unadjusted trends, but the initial crossover date was delayed slightly until April 11 (adjusted RR = 1.08, 95% PI: 1.01, 1.14). The RRs from June to mid-August were significantly positive, but the values were attenuated relative to the unadjusted model, achieving a maximum of 1.99 (95% PI: 1.86, 2.13) on June 21. This attenuation suggests that adjustment accounted for some of the differential effect between upper and lower SVI counties. Of note, by August 31, there was second crossover event in which the lower quartile surpassed the upper quartile in per capita cases (RR: 0.74, 95% PI: 0.66, 0.87). We found similar trends for the death rate models Figures S8A-B). Here, the initial crossover date was delayed until June 6 (RR = 1.34, 95% PI: 1.00, 1.28) and the RRs for June-August were attenuated, with a maximum of 1.62 (95% PI: 1.37, 1.85) on July 28. In general, the same patterns emerged for the SVI themes: the initial crossovers were delayed, the RRs were attenuated during the summer months, and by late August, the lower quartile matched or superseded the upper quartile in per capita trends (Figures S9–S16). For the Race/Ethnicity/Language theme, the incidence rate on August 31 for the upper quartile was approximately half that for the lower quartile (RR = 0.52, 95% PI: 0.47, 0.58). Thus, controlling for variable such as rurality, health, and PM2.5, appeared to account in part for the differences between upper and lower quartiles observed in the unadjusted analyses.

There were also several significant associations among the adjustment variables (Table 1). Percent rural and percent smoking were negatively associated with COVID-19 cases (percent rural RR = 0.78, 95% PI: 0.76, 0.79; percent smoking RR = 0.81, 95% PI: 0.79, 0.83), as well as deaths (percent rural RR = 0.77, 95% PI: 0.75, 0.80; percent smoking RR = 0.76, 95% PI: 0.73, 0.79). In contrast, percent in fair or poor health, average PM2.5, and state proportion tested were positively associated with both cases (percent poor/fair health RR = 1.48, 95% PI: 1.45, 1.51; PM2.5 RR = 1.26, 95% PI: 1.24, 1.28; proportion tested RR = 1.08, 95% PI: 1.06, 1.10) and deaths (percent poor/fair health RR = 1.81, 95% PI: 1.74, 1.88; PM2.5 RR = 1.31, 95% PI: 1.27, 1.45; proportion tested RR = 1.13, 95% PI: 1.10, 1.16). Number of primary care physicians per 100,000 was associated with fewer cases (RR = 0.97, 95% PI: 0.95, 0.99), but was not associated with deaths (RR = 1.00, 95% PI: 0.97, 1.03).

**Table 1.** Adjusted risk ratios (RRs) and 95% posterior intervals (PIs) for the adjustment variables in adjusted analysis

| Model Outcome | Variable | Risk Ratio (95% PI) |
| --- | --- | --- |
| Incident Cases | % of county designated rural<sup>*</sup> | 0.78 (0.76, 0.79) |
|  | % fair or poor health in county<sup>*</sup> | 1.48 (1.45, 1.51) |
|  | % adult smokers in county<sup>*</sup> | 0.81 (0.79, 0.83) |
|  | Average daily PM<sub>2.5</sub> for county<sup>*</sup> | 1.26 (1.24, 1.28) |
|  | Proportion tested in the state<sup>†</sup> | 1.08 (1.06, 1.10) |
|  | # primary care physicians per 100,000 in county<sup>*</sup> | 0.97 (0.95, 0.99) |
| Deaths | % of county designated rural<sup>*</sup> | 0.77 (0.75, 0.80) |
|  | % fair or poor health in county<sup>*</sup> | 1.81 (1.74, 1.88) |
|  | % adult smokers in county<sup>*</sup> | 0.76 (0.73, 0.79) |
|  | Average daily PM<sub>2.5</sub> for county<sup>*</sup> | 1.31 (1.27, 1.45) |
|  | Proportion tested in the state<sup>†</sup> | 1.13 (1.10, 1.16) |
|  | # primary care physicians per 100,000 in county<sup>*</sup> | 1.00 (0.97, 1.03) |

<sup>\*</sup> From Robert Wood Johnson Foundation's 2019 County Health Rankings & Roadmaps: Rankings Data & Documentation
<sup>†</sup> As of August 31, 2020

Finally, Tables S1-S4 in the Supporting Information present the top 10 counties with the highest average incidence (Tables S1 and S2) and death rates (Tables S3 and S4) from the unadjusted and adjusted models for the week of August 24 – 31, 2020. There was substantial overlap in the unadjusted and adjusted rankings, with the unadjusted models ranking at the top southeastern counties like Wayne, Tennessee, and Chattahoochee, Georgia, while the adjusted models pick up on emerging trends in the northern Midwest and Mountain states, including Rosebud, Montana, and Custer, South Dakota.

## Discussion

In this study, we hypothesized that counties with greater vulnerability would have higher COVID-19 incidence and death rates compared to less vulnerable counties and that this disparity would widen over time. Overall, the incidence and death rates increased for both the more and less socially vulnerable counties from March 1 to August 31, but the rates of increase varied depending on the time period. For some SVI themes, we found that less vulnerable counties, such as Nassau County, New York, had slightly higher average incidence and death rates early in the pandemic compared to more vulnerable counties, such as Brooks County, Texas. However, by April and May 2020, the trends crossed, with the most vulnerable counties experiencing, on average, substantially higher burden from the disease compared to less vulnerable counties. This crossover effect could be the result of state re-openings, which may have disproportionately impacted more vulnerable counties. Crossover effects were observed for overall SVI (cases), as well as Socioeconomic Status (cases and deaths) and, most notably, Household Composition (cases and deaths), where the crossover date was delayed until mid-May. This theme represents elderly and individuals with disabilities, and may reflect early outbreaks at long-term care facilities in lower vulnerability areas such as King County, Washington [38]. For most SVI themes, incident cases and deaths among the upper quartile counties outpaced those in the lower quartile through July, with the most notable disparity occurring for the Race/Ethnicity/Language theme. In many cases, the RRs declined in early August, as the lower quartile counties kept pace with those in the upper quartiles. For some SVI themes, including Race/Ethnicity/Language, we observed a second crossover event in late August, when the lower quartile surpassed the upper quartile in per capita cases and deaths. These patterns held up after adjustment and in sensitivity analyses using Johns Hopkins data. In fact, to our knowledge, this is the first study to track COVID-19 trends across multiple data repositories.

Our findings are generally consistent with the study by Khazanchi et al. examining data up to April 19, 2020, which found that counties in the top quartile of overall SVI had higher incidence and death rates compared to those in the lower quartile [19]. As in that study, we found the strongest disparity for the Race/Ethnicity/Language theme. However, Khazanchi et al. found no association with Household Composition, whereas we found that the lower quartile had higher rates of cases and deaths during this period. This may be due to the fact that the authors looked at cumulative cases through April 19, whereas we examined daily incidence. Moreover, through our longitudinal analysis, we observed that overall SVI, Socioeconomic Status and Housing Composition had negative RRs for much of March and early April. This highlights the benefit of the longitudinal approach: it provides a comprehensive picture of the evolving relationship between SVI and COVID-19, rather than a momentary snapshot.

Our findings may also explain inconsistent findings in two other studies. As in our study, Karaye et al. found that overall SVI and Race/Ethnicity/Language were associated with increased COVID-19 incidence through May 12, 2020 [17]. However, they found no association between Socioeconomic Status and incident cases, whereas Household Composition and Housing/Transportation had an inverse relationship. Our results place these findings in temporal context. In particular, we found a delayed crossover effect for Household Composition, with RRs below or near 1.00 through mid-May. In particular, on May 12, we found a null association for Household Composition (RR = 0.98, 95% PI: 0.93, 1.04) in agreement with Karaye et al; however, just days later, on May 16, we found a significant positive association (RR = 1.06, 95% PI: 1.01, 1.11). Nayak et al., meanwhile, found no association between overall SVI and cumulative COVID-19 incidence on April 4, 2020 [18]. According to our results, however, this was close to the crossover dates of March 30 (unadjusted analysis) and April 11 (adjusted analysis), a period in which the disparity between high and low SVI counties hovered near 1.00. By mid-April, we observed consistent positive associations between overall SVI and both cases and deaths. Additionally, Nayak and colleagues found that the Race/Ethnicity/Language and Housing/Transportation themes were positively associated with incident cases, but Household Composition was not. However, we found that the RRs for Household Composition varied over time. Again, these results highlight the need to consider both temporal and spatial variability when attempting to fully understand, in real time, the impact of the pandemic on populations with different vulnerability profiles.

Several covariates from our adjusted model were significantly associated COVID-19 cases and deaths. We found that rurality was associated with fewer cases and deaths, consistent with a prior study [19]. In contrast, percentage in poor or fair health was positively associated with both cases and deaths. This supports results from a recent study that found that patients with COVID-19 with cardiovascular disease, hypertension, diabetes mellitus, congestive heart failure, chronic kidney disease, and cancer had a higher risk of mortality, compared to patients with COVID-19 without these comorbidities [30]. Moreover, as in prior studies [17, 28], we found that average PM2.5 was positively associated with both cases and deaths. Increased state-level testing was also associated with higher rates of COVID-19 cases and deaths, likely due to heightened surveillance. Contrary to our expectation, we found a significant inverse association between percentage of adult smokers and COVID-19 cases deaths. Our aggregated, county-level findings support recent individual-level studies suggesting that nicotine may have a protective effect on COVID-19 [39, 40]. Finally, the number of primary care physicians per capita was associated with lower incidence, but there was no association with deaths.

More generally, our results suggest a dynamic impact of COVID-19 on socially vulnerable communities. Contrary to expectation, we found that COVID-19 disproportionately impacted less vulnerable counties early in the pandemic, before spreading to more vulnerable areas in May-July. This shift could reflect local and state policy decisions, such as early re-openings in states like Georgia with a high percentage of vulnerable counties [41, 42]. By August, however, the least vulnerable counties began to keep pace with the more vulnerable counties, suggesting that the impact of COVID-19 is not static, but can migrate from less vulnerable counties to more vulnerable ones and back again over time. These results highlight the need for communities, even less vulnerable ones, to continue to monitor the spread of the disease, maintain adequate health care resources, and implement local social distancing measures.

Our analysis sheds light on the community-level burden of COVID-19 as measured by population-adjusted incidence and death. This information can be used to inform policy decisions related to COVID-19 and future pandemics. For example, our model can be used to detect county-specific spikes, plateaus, and troughs that reflect outbreaks at nursing homes or correctional facilities, as well as the impact of in policy changes, such as stay at home orders and statewide re-openings of public spaces and local businesses, or the return to schools and universities. Moreover, the model provides for accurate prediction of COVID-19 trends for individual counties, allowing health officials to target intervention. By monitoring changes in temporal trends, local policymakers can mobilize resources to minimize imminent outbreaks.

There are also limitations to this analysis. First, our analysis is largely descriptive with the goal of generating hypotheses to inform policy and guide future research. For example, future studies might review the policy actions that gave rise to the crossover effects we observed early and late in the pandemic for several of the SVI themes. Second, we used county-level SVI data from 2018. It is possible that social vulnerability factors may have changed between 2018 and 2020, but we used the most recent SVI data available from CDC. Third, we downloaded several of the adjustment variables from the Robert Wood Johnson Foundation’s 2019 County Health Rankings & Roadmaps database, which may not be the most current source for variables such as PM2.5. Fourth, it was challenging to model deaths because most counties reported no deaths on any given day. Future studies could employ zero-inflated models to better account for this aspect of the data [43–45]. Future work could also examine temporal trends in locations of correctional facilities, long-term care facilities, nursing homes, Indian reservations and Tribal lands, and other places with high rates of infection [46–49]. Finally, we examined trends in the US only; future work might replicate our study in developing countries or those with emerging outbreaks.

Examining the impact of COVID-19 on vulnerable communities in the US is of growing importance [15, 50]. Mounting evidence suggests that social determinants of health and community contextual factors contribute to disparities in both COVID-19 incident cases and deaths [2, 3, 6, 51]. It is therefore critically important to monitor and protect vulnerable populations as the pandemic continues to unfold.

## Data Availability

Data used for this study are publicly available. Computing code is available upon request from the corresponding author.

## Abbreviations

CDC: Centers for Disease Control and Prevention
COVID-19: coronavirus disease 2019
PI: posterior interval
RR: risk ratio
SARS-CoV-2: severe acute respiratory syndrome coronavirus 2
SVI: Social Vulnerability Index
US: United States

